# What content and methods are effective in developing cognitive enhancement programs for older adults with mild cognitive impairment? A cross-sectional study

**DOI:** 10.64898/2025.12.20.25342600

**Authors:** Younkyoung Kim, Saeryun Kim, Keiko Asami

## Abstract

Early intervention in older adults with mild cognitive impairment (MCI) is effective in maintaining and enhancing cognitive function. To maximize the effectiveness of such cognitive enhancement programs, it is essential to adequately reflect the characteristics and needs of the target population. This study aimed to assess the basic competencies of older adults with MCI, explore their needs and memorable experiences, and identify appropriate content and delivery strategies for cognitive enhancement programs. A cross-sectional design was employed, combining survey data and in-depth interviews. A total of 130 community-dwelling older adults with MCI living in both urban and rural areas of South Korea participated. Quantitative data included sociodemographic characteristics, cognitive function, health literacy, basic physical capability, and mobile device proficiency, and were analyzed using descriptive statistics, independent t-tests, and chi-square tests. Qualitative interview data were analyzed using inductive content analysis. The findings showed that participants had generally low levels of education, limited health literacy, and low mobile device proficiency, despite a high rate of smartphone ownership. Interview results revealed a strong demand for cognitive enhancement programs and suggested that participants preferred content related to family, recreational activities, career, academics, and Korean traditional holidays. These results underscore the need for cognitive enhancement programs that are tailored to the specific characteristics of community-dwelling older adults with MCI. Effective programs should take into account their lower levels of education, literacy, and mobile device proficiency, and incorporate content that reflects their needs and memorable experiences.

## 1. BACKGROUND

Mild cognitive impairment (MCI) represents the transitional zone between normal age-related cognitive decline and early dementia (Albert at al., 2011; Roberts & Knopman, 2013). Individuals aged 65 and older who have MCI exhibit a dementia incidence rate of 241.3 per 1,000 person-years, significantly higher than that of older adults without cognitive impairment (Zhang et al., 2021). Pharmacological treatments have not demonstrated sufficient effectiveness in preventing cognitive deterioration or further decline (Fink et al., 2020), and no medications have been approved to prevent the progression from MCI to dementia beyond 36 months (Alzheimer’s Association, 2019; Petersen 2016). However, early intervention can help maintain or improve cognitive abilities (Rodakowski et al., 2015); delaying dementia progression by five years could potentially reduce dementia cases by up to 57% (Sperling et al., 2011). As such, cognitive enhancement programs hold promise in dementia prevention for individuals with MCI.

Several programs aimed at improving cognitive abilities in older adults with MCI have been reported. Cognitive enhancement programs targeting MCI generally include cognitive training and rehabilitation programs (focusing on verbal memory, executive function, attention, processing speed, etc.), physical activity programs (such as aerobic exercise, muscle-strengthening training, Tai chi, balance and core resistance training, and walking), or multidomain programs combining these components (Salzman et al., 2022). These programs have been delivered using various methods, including interactive workbooks, group social activities like tea-drinking meetings, gaming, and digital device-based interventions (Kim et al., 2023; Chalfont et al., 2021).

Older adults with MCI, in comparison to older adults in general, often demonstrate particular characteristics associated with cognitive decline. Individuals with MCI often have lower health literacy than those with normal cognitive function (Liu et al., 2019), and cognitive decline may impair their ability to perform activities of daily living such as shopping and banking (Rodakowski et al., 2014). This decline may lead to decreased self-care performance (Ellis et al., 2019). These changes in older adults with MCI can make it difficult for them to consistently participate in cognitive enhancement programs. Therefore, effective cognitive enhancement programs for people with MCI must be replicable, adaptable, and tailored (Ellis et al., 2019) and programs should require minimal resources and be seamlessly integrated into existing medical and community services (Kinsella et al., 2016). Consequently, meticulous examination of these programs is essential to ensure that they align with the characteristics of older adults with MCI, as well as their preferences and their specific requirements for cognitive enhancement programs. Furthermore, not only quantitative data but also detailed qualitative data are needed to identify the actual needs of older adults with MCI. This information is necessary to develop specific strategies and content that address their needs, thus facilitating the effective application of cognitive enhancement programs.

The purpose of this study was to assess cognitive abilities, physical characteristics, health literacy, digital literacy, and both needs and memorable experiences related to cognitive enhancement programs among community-dwelling older adults with MCI. The results of this study can provide a basis for understanding the characteristics and basic competencies of older adults with MCI, and inform the development of appropriate program content and delivery methods, taking into account their needs and memorable experiences.

## 2. METHODS

### 2.1. Research design and setting

This study employed a two-method approach to explore the basic competencies and characteristics of older adults with MCI, along with their needs and memorable experiences regarding the content and methods used in cognitive enhancement programs conducted in South Korea between October 2022 and March 2023 Participants were recruited through national dementia care and older-adult welfare centers located in urban and rural areas of South Korea. The inclusion criteria for the study were: (1) aged 65 years or older; (2) physician-diagnosed MCI; (3) a Korean version of the Montreal Cognitive Assessment (K-MoCA) score of 17 - 23; and (4) proficiency in reading and writing Korean. Exclusion criteria included: (1) any form of dementia as defined by Diagnostic and Statistical Manual of Mental Disorders (DSM) −5 and International Classification of Diseases (ICD) −10 guidelines; (2) communication difficulties; and (3) cognitive impairment resulting from brain injury or stroke. Data were collected through surveys and interviews in a private setting. Interviews were audio-recorded with participants’ consent, and data were gathered using audio recordings and notes.

The sample size was calculated using G*Power 3.1, which indicated a required sample size of 128 participants (64 per region). Accounting for an anticipated dropout rate of 9% (Jeoung et al., 2022), 142 participants were recruited. Following initial screening, 12 individuals who did not meet the K-MoCA score criteria, were unable to write, or exhibited significant communication difficulties were excluded, resulting in 130 eligible participants (65 from urban areas and 65 from rural areas).

### 2.2. Variables and measurement

Data were collected through structured questionnaires and semi-structured individual interviews. The structured questionnaire assessed general characteristics (sex, age, educational level, employment, monthly household income, smartphone user, household composition, and primary caregiver), health-related characteristics (subjective health status, use of assistive devices, medical history, exercise), and characteristics related to participation in cognitive enhancement programs. Additional structured questionnaire items are described below.

#### Cognitive function

Cognitive function was assessed using the K-MoCA (Kang et al., 2009), initially developed by Nasreddine et al. (2005). The K-MoCA evaluates visuospatial/executive function, naming, memory, attention, language, abstraction, delayed recall, and orientation, yielding a total possible score of 30 points. Scores between 17 and 23 indicate MCI (Kim et al., 2023).

#### Health literacy

Health literacy was measured with the Korean version of the Newest Vital Sign (NVS) (Kim, 2011), originally developed by Weiss et al. (Weiss et al., 2005). This instrument consists of six items, each scoring 1 for a correct answer and 0 for an incorrect answer, producing a total score ranging from 0 to 6. Scores of 0–1 indicate a high likelihood of limited literacy, scores of 2–3 suggest possible limited literacy, and scores of 4–6 indicate adequate literacy. Reliability during its development was reflected by a Cronbach’s α of .76, and in this study, KR-20 was .60.

#### Basic physical capability

Basic physical capability was assessed using the Korean Version of the Basic Physical Capability Scale (BPCS-K) (Song & Hong, 2020), initially developed by Resnick et al. (2014). The BPCS-K comprises 16 items across five domains, with higher scores indicating better physical capability. Reliability of this instrument was KR-20 = .91 at the time of translation and .76 in this study.

#### Mobile device proficiency

Mobile device proficiency was evaluated using the Mobile Device Proficiency Questionnaire-16 (MDPQ-16), which was developed by Roque & Boot (2018) and translated into Korean by our research team according to World Health Organization guidelines (2018). The translation was reviewed for face validity by seven older adults and for content validity by five experts (CVI = 0.99). Based on expert feedback, an additional item on mobile messenger use (e.g., KakaoTalk) was included, creating a total of 18 items across several domains: mobile basics, communication, data storage, internet use, calendar management, entertainment, privacy settings, and troubleshooting. Participants rated their proficiency on each item using a 5-point Likert scale, with higher scores indicating greater proficiency. Reliability was Cronbach’s α = .96 in the original study (Roque & Boot, 2018) and Cronbach’s α = .95 in the current study.

#### Semi-structured interview form

The semi-structured interview questions were developed to assess participants’ needs related to cognitive enhancement programs and to generate program content. Questions underwent content validation by five nursing professors, resulting in a CVI of 1.0. The questions were designed to include topics participants remember, talk about, or wish to share from their whole lives. Key questions included: “What is your experience with participating in cognitive enhancement programs?” and “What are the things in your life that you want to remember and come back to?”

### 2.3. Statistical analysis

Quantitative data were analyzed using SPSS Statistics for Windows, version 25.0 (IBM Corp., Armonk, NY, USA) with two-tailed tests at a significance level of .05. Descriptive statistics, independent t-tests, and chi-square tests were conducted to compare characteristics of MCI participants between urban and rural areas.

Qualitative interview data were analyzed using inductive content analysis (White & Marsh, 2006), during which researchers identified themes and patterns by reviewing transcripts. Revisions continued until a consensus was reached.

### 2.4. Ethical consideration

This study was approved by the Institutional Review Board of the primary researcher’s institution (No. 1040198-220819-HR-104-02). All participants provided written and oral information about the study, including the purpose and procedures of this study, right to withdraw and the guarantee of anonymity. All participants signed voluntary a statement of an Informed Consent Form prior to participation of this study.

## 3. RESULTS

### 3.1. Quantitative results

In this study, 130 participants were included, of whom 78 (60.0%) were women, with an average age of 79.36 ± 4.90 years. The proportion of elementary school graduates was highest overall. However, there was a statistically significant difference in educational level between the two regions (χ² = 17.05, *p* = .001). Among participants, 83.1% were unemployed, and 77.7% reported a monthly household income of less than $720. Regarding household composition, participants living with a spouse only represented the highest percentage (47.7%), although household composition showed a significant difference between the two regions (χ^2^ = 7.70, *p* = .021). A total of 34.6% reported previous participation in a cognitive enhancement program, and willingness to join future programs was expressed by 50.8% in urban and 56.9% in rural areas (see Table 1).

**Table 1.**
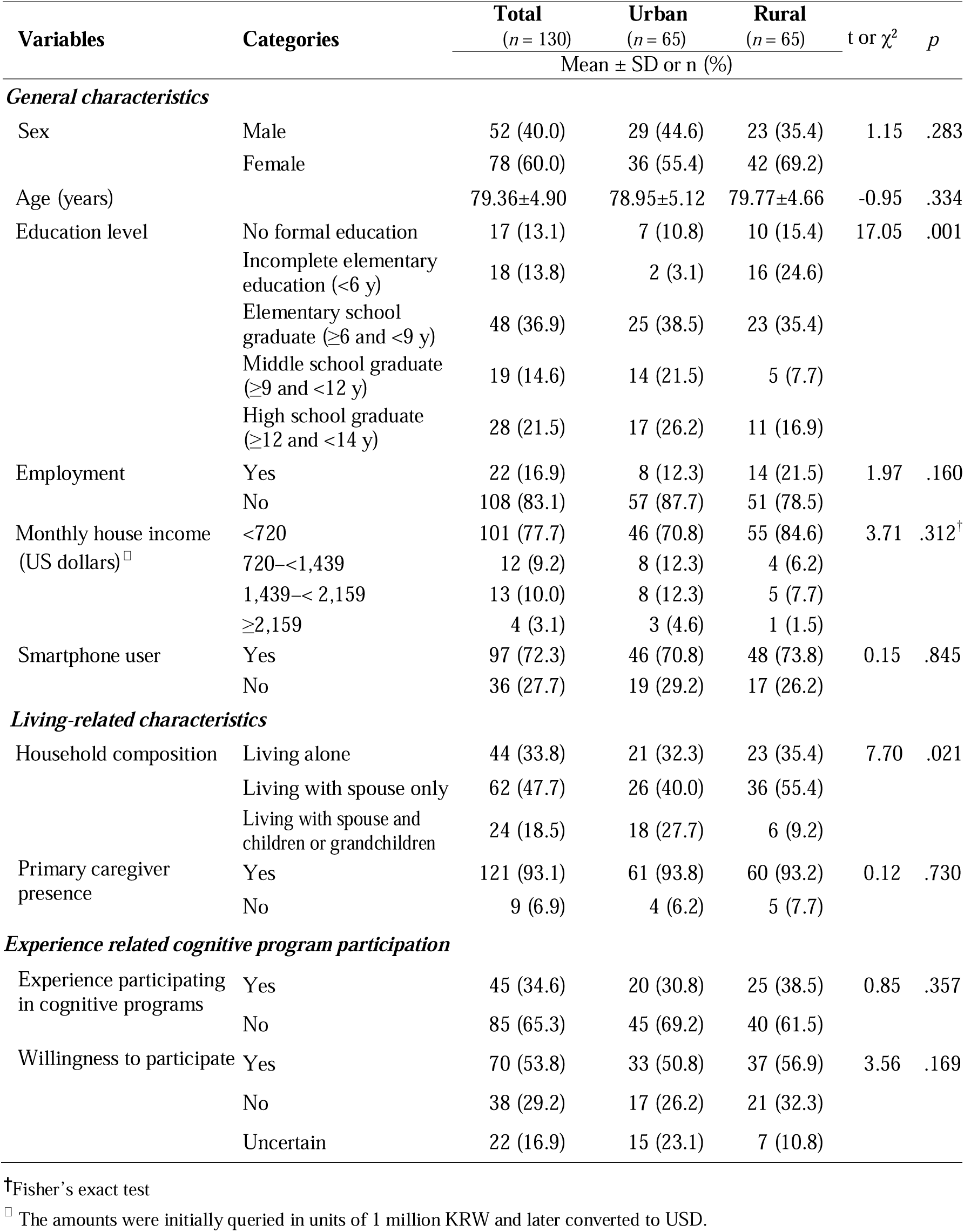
General characteristics of participants (N = 130)

| Variables | Categories | Total | Urban | Rural | t or $\chi^2$ | p |
| --- | --- | --- | --- | --- | --- | --- |
|  |  | (n = 130) | (n = 65) | (n = 65) |  |  |
| Mean ± SD or n (%) | | | | | | |
| *General characteristics* |  |  |  |  |  |  |
| Sex | Male | 52 (40.0) | 29 (44.6) | 23 (35.4) | 1.15 | .283 |
|  | Female | 78 (60.0) | 36 (55.4) | 42 (69.2) |  |  |
| Age (years) | | 79.36±4.90 | 78.95±5.12 | 79.77±4.66 | -0.95 | .334 |
| Education level | No formal education | 17 (13.1) | 7 (10.8) | 10 (15.4) | 17.05 | .001 |
|  | Incomplete elementary education (<6 y) | 18 (13.8) | 2 (3.1) | 16 (24.6) |  |  |
| | Elementary school graduate (≥6 and <9 y) | 48 (36.9) | 25 (38.5) | 23 (35.4) | | |
| | Middle school graduate (≥9 and <12 y) | 19 (14.6) | 14 (21.5) | 5 (7.7) | | |
| | High school graduate (≥12 and <14 y) | 28 (21.5) | 17 (26.2) | 11 (16.9) | | |
| Employment | Yes | 22 (16.9) | 8 (12.3) | 14 (21.5) | 1.97 | .160 |
|  | No | 108 (83.1) | 57 (87.7) | 51 (78.5) |  |  |
| Monthly house income (US dollars)<sup>□</sup> | <720 | 101 (77.7) | 46 (70.8) | 55 (84.6) | 3.71 | .312<sup>†</sup> |
|  | 720–<1,439 | 12 (9.2) | 8 (12.3) | 4 (6.2) |  |  |
|  | 1,439–<2,159 | 13 (10.0) | 8 (12.3) | 5 (7.7) |  |  |
| | ≥2,159 | 4 (3.1) | 3 (4.6) | 1 (1.5) | | |
| Smartphone user | Yes | 97 (72.3) | 46 (70.8) | 48 (73.8) | 0.15 | .845 |
|  | No | 36 (27.7) | 19 (29.2) | 17 (26.2) |  |  |
| *Living-related characteristics* |  |  |  |  |  |  |
| Household composition | Living alone | 44 (33.8) | 21 (32.3) | 23 (35.4) | 7.70 | .021 |
|  | Living with spouse only | 62 (47.7) | 26 (40.0) | 36 (55.4) |  |  |
|  | Living with spouse and children or grandchildren | 24 (18.5) | 18 (27.7) | 6 (9.2) |  |  |
| Primary caregiver presence | Yes | 121 (93.1) | 61 (93.8) | 60 (93.2) | 0.12 | .730 |
|  | No | 9 (6.9) | 4 (6.2) | 5 (7.7) |  |  |
| *Experience related cognitive program participation* |  |  |  |  |  |  |
| Experience participating in cognitive programs | Yes | 45 (34.6) | 20 (30.8) | 25 (38.5) | 0.85 | .357 |
|  | No | 85 (65.3) | 45 (69.2) | 40 (61.5) |  |  |
| Willingness to participate | Yes | 70 (53.8) | 33 (50.8) | 37 (56.9) | 3.56 | .169 |
|  | No | 38 (29.2) | 17 (26.2) | 21 (32.3) |  |  |
|  | Uncertain | 22 (16.9) | 15 (23.1) | 7 (10.8) |  |  |

<sup>†</sup>Fisher's exact test<sup>□</sup> The amounts were initially queried in units of 1 million KRW and later converted to USD.

Table 2 presents the health-related characteristics of participants. A total of 46.2% of participants rated their subjective health status as fair. Among those using assistive devices (65.4%), 42.0% used eyeglasses and 24.1% used a cane. Additionally, 93.8% reported having diagnosed diseases. Overall, 56.9% reported not engaging in muscle-strengthening exercises, with a significant difference in exercise performance between the urban and rural groups (χ^2^ = 15.18, *p* < .001). The average weekly duration of muscle-strengthening exercises differed significantly between the two groups (t = 3.87, *p* < .001), with walking being the most common type of exercise (41.4%).

**Table 2.**
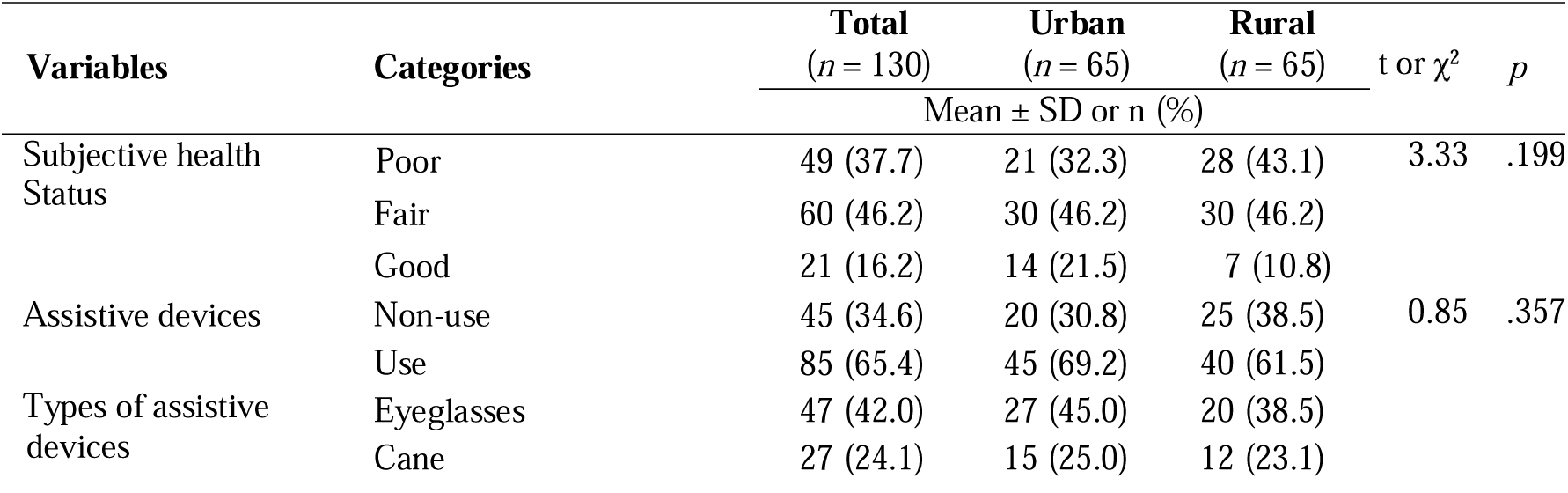
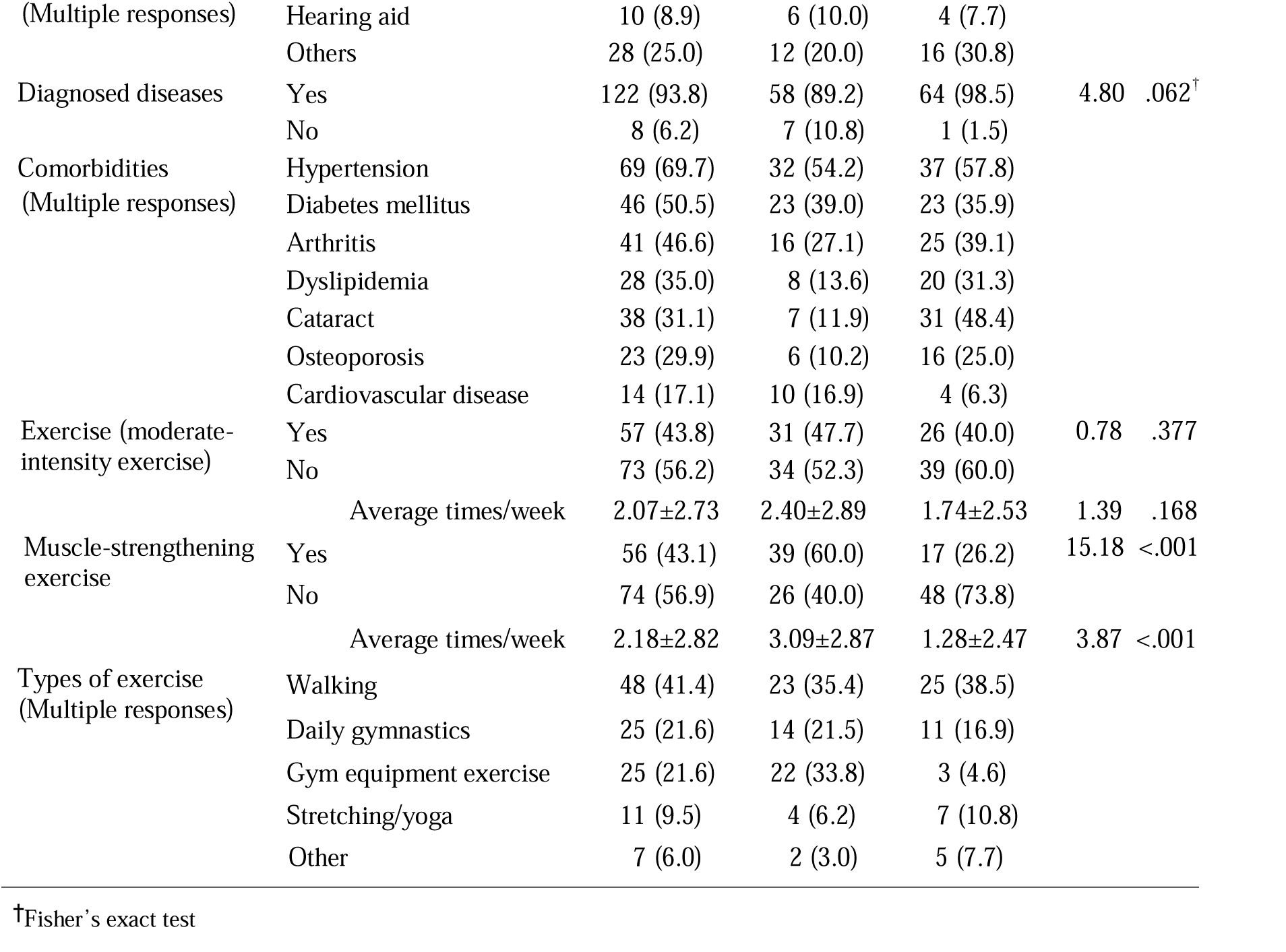
Health-related characteristics of participants (N = 130)

Cognitive function scores averaged 18.38 ± 2.01 (t = 1.05, *p* = .296) and basic physical capability averaged 14.89 ± 1.70 (t = −1.46, *p* = .149). The average health literacy score was 1.06 ± 1.15 (t = −0.92, *p* = .361), indicating a high likelihood of limited literacy; no statistically significant differences emerged between urban and rural participants. In contrast, mobile device proficiency scores differed significantly between urban and rural groups (t = 2.27, *p* = .025) (see Table 3).

**Table 3.** Characteristics of main variables (N = 130)

| Variables | Total<br>(*n* = 130) | Urban<br>(*n* = 65) | Rural<br>(*n* = 65) | t | *p* |
| --- | --- | --- | --- | --- | --- |
|  | Mean ± SD |  |  |  |  |
| K-MoCA | 18.38±2.01 | 18.57±1.83 | 18.20±2.17 | 1.05 | .296 |
| Health literacy | 1.06±1.15 | 0.98±1.16 | 1.15±1.13 | -0.92 | .361 |
| BPCS-K | 14.89±1.70 | 14.68±2.08 | 15.11±1.17 | -1.46 | .149 |
| Mobile device proficiency | 12.72±6.41 | 13.97±7.80 | 11.46±4.34 | 2.27 | .025 |

*Note:* K-MoCA: Korean version of the Montreal Cognitive Assessment, BPCS-K: Korean Version of the Basic Physical Capability Scale

### 3.2. Qualitative results

Table 4 summarizes interview findings regarding the needs for participation in cognitive enhancement programs. Older adults with MCI expressed willingness to participate in future cognitive enhancement programs due to their pursuit of an active life, enjoyment of engagement, and expectations of dementia prevention. Conversely, reluctance to engage in cognitive enhancement programs was related to physical factors, such as inconvenience due to physical distance, limited time available because of economic activities, and limitations in participating in group activities due to physical difficulties, as well as emotional factors, including lack of confidence in their ability to complete the program compared to others and fear of private information exposure.

**Table 4.** Needs for participation in cognitive enhancement programs (N = 130)

| Categories | Sub-categories | Quotes |
| --- | --- | --- |
| Willingness to participate | Pursuing an active life | *As I get older, I feel I shouldn't stay inactive, so I decided to join the program. (ID 4)* |
|  | Joy in engagement | *I'll participate because it brings me joy. (ID 10)* |
|  | Expectations for dementia prevention | *I should join as a preventive measure against dementia. (ID 98)*<br>*If it can help improve my memory, I should definitely participate. (ID 116)* |
| Reluctance to engage | Inconvenience due to physical distance | *I can't participate because traveling long distances is uncomfortable for me. It would be more convenient if it were closer. (ID 1)* |
|  | Lack of time for participation in economic activities | *I can't participate because my schedule conflicts with my public work responsibilities. (ID 127)* |
|  | Lack of confidence in completing the program like others | *I don't want to do it because I lack confidence in my ability. (ID 6)* |
|  | Limitations on group activities due to physical issues | *I don't want to join group activities because my physical discomfort makes it hard to keep up with others. (ID 5)*<br>*I want to participate, but my physical discomfort and lack of confidence are holding me back. (ID 39)* |
|  | Fear of exposing private parts | *I don't want to participate because I prefer to keep my personal life private. (ID 67)* |

The analysis of interview data yielded 18 subcategories grouped into five categories (see Table 5). The “Family” category revealed memories involving the family of origin and the joy derived from raising children. The “Recreational Activities” category highlighted games and activities with friends, craftwork, and longing for physical activities. The “Career” category encompassed professional achievements, regrets, and aspirations. The “Academics” category reflected academic success, feelings of loss from leaving school, and desire to continue learning. Lastly, the “Korean Traditional Holidays” category captured memories of holiday customs and the richness of traditional experiences.

**Table 5.** Structured qualitative data for organizing cognitive program content.

| Categories | Sub-categories | Quotes | Application Example |
| --- | --- | --- | --- |
| Family | Efforts to help family during childhood | *I was the oldest daughter, so I helped my mom with her business and took care of my younger siblings at home. (ID 49)*<br>*When I was a kid, it was hard because we didn't really have anything to eat... I was very young but I used to go and get pebbles for flour. (ID 70)* | ☑ Content: Writing or drawing for myself who overcame difficult times (executive function) |
|  | Love and care received from parents | *My mom sewed a beautiful outfit and made me wear it. (ID 10)*<br>*When it was just the three of us, Mom, Dad and I, we'd have a campfire at night in the mountains, and Dad would tell me the constellations, and we'd have fun. (ID 104)* | ☑ Content: Hidden picture search in a birthday party illustration (visuospatial function) |
|  | Cherished memories with family | *I loved traveling with my family, seeing flowers, seeing water, seeing rivers. (ID 3)*<br>*My brother and I used to travel by train with a phonograph. (ID 10)* | ☑ Content: Matching seasonal changes (recall, orientation) |
|  | Happiness in a child's growth and success | *My eldest daughter majored in English at university, and when she graduated, she got a job in a big company as a secretary interpreting for the chairman, and she was very proud of herself. She did well in her studies. (ID 58)*<br>*My favorite thing is seeing my kids grow up healthy and thriving. (ID 108)* | ☑ Content: Designing a calendar and marking your child's birthday (recall, orientation, executive function) |
|  | Joy and self-worth from cooking for family | *My children say my mom's side dishes are the best. Everyone says I'm a good cook. I'm going to make scallion kimchi to share with my children. (ID 82)*<br>*I'm happy to serve my kids kimchi or something. (ID 74)* | ☑ Content: Selecting the ingredients and ordering the steps for making kimchi (association, executive function) |
|  | Wishes for a family trip | *I want to travel abroad to travel abroad with my family someday. (ID 15)*<br>*It's nice to get together with the family and go on a trip or something. (ID 55)* | ☑ Content: Planning a trip with family (executive function) |
| Recreational Activities | The joy of various childhood games | *When I was a kid, we used to go sledding in the winter on sleds made of wooden planks and splashing in the summer. (ID 37)*<br>*When I was a child, I played marbles, territory games, jackstone games, and shuttlecock kicking. (ID 83)* | ☑ Content: Playing a marble game and calculating each score (physical exercise, calculation) |
|  | Nature-based gathering activities | *I would go to the river with my friends and catch fish, go to the fields and pick ginseng to eat and play. (ID 9)*<br>*It reminds me of going to the beach with my friends and playing with mudskippers and whelks. (ID 25)* | ☑ Content: Identifying farm animals and counting their numbers (calculation, executive function) |
|  | The happiness of traveling with friends | *My favorite time was when I traveled to famous Mountain and Island with my friends. (ID 62)*<br>*My happiest moments were going to the mountains and hanging out with my friends. (ID 101)* | ☑ Content: Checking the map and listing major travel destinations in country (recall, visuospatial function, language) |
|  | Accomplishment in craft activities | *I like to knit socks or something like that. I feel good when I make something (ID 71).*<br>*I'm good at making baskets with my own hands. (ID 97)* | ☑ Content: Cutting out shapes from paper and creating animal figures (visuospatial function, executive function) |
|  | Longing for physical activities | *I can't play tennis anymore because I've been away from it for so long. (ID 20)*<br>*When I see young people doing sports, gymnastics, and stuff like that, I think, 'How do they do it so well?' I can't do it now, so when I see that, I'm envious. (ID 72)* | ☑ Content: Calisthenics (physical exercise) |
| Career | Pride in professional achievements | *The best times were when I was young, doing well and making a lot of money. (ID 35)*<br>*When I worked in a skilled job, I studied the design and made the clothes to fit the customer, and when the customer wore the clothes and said, "Thank you, Mr. Boss," I felt very happy. (ID 18)* | ☑ Content: Matching uniforms to the appropriate professions (orientation, recall) |
|  | Unfulfilled aspirations in career | *I used to be told I had a beautiful voice, and I wanted to be a voice actor, but I couldn't. (ID 46)*<br>*I wish I could have continued with the transportation business, but I was broke and couldn't. (ID 52)* | ☑ Content: Drawing a picture of a wish (executive function) |
| Academics | Positive memories from school successes | *I was happiest in elementary school when I won honors. (ID 28)*<br>*I won a prize for running well at a sports day, which was nice. (ID 73)* | ☑ Methods: Issuing a certificate of completion after participating in a cognitive enhancement program |
|  | Feeling of loss from leaving school | *I didn't go to school much in the past, so if I were younger, I'd like to go back to school more. It's the one thing I really regret not learning. (ID 74)*<br>*I remember when I was a kid, only my older brother went to school and I was at home watching him go to school, and it was the most heartbreaking thing. (ID 102)* | ☑ Methods: Using name badges, attendance check, and praise stickers or stamps in school lessons |
|  | Desire for education | *I want to study, but my mother always told me to do chores because she was a farmer, so I didn't study. (ID 100)*<br>*I wish I could have gone to school like everyone else, that's what I envy the most. (ID 130)* | ☑ Methods: Providing learning workbooks similar to school homework |
|  | Need for learning digital devices | *I could really use to learn how to use the Computer. (ID 2)*<br>*I would like to learn how to use my mobile phone (ID 39)* | ☑ Methods: Cognitive enhancement program using digital devices |
| Korean traditional holidays | Enjoying traditional holiday activities | *I've gone on holiday parties, played moonwalking, and stayed up until dawn with my friends. (ID 42)*<br>*I can't tell you how many holidays I've waited for to wear a Korean traditional clothing skirt, a Traditional Upper Garment, and so on. (ID 15)* | ☐ Content: Playing traditional holiday games while following the rules (physical activity, calculation) |
|  | A feast of holiday dishes | *It's really good for holidays. I used to eat rice cake for Korean thanksgiving day, honey cookies and crispy rice snack for Lunar New Year. (ID 3)*<br>*During the holidays, it's nice to get together with family and cook a lot of food for each other. (ID 4)* | ☐ Content: Matching different foods to their respective holidays and writing their names (association, language) |
|  | Family togetherness during holidays | *On holidays, my children bring microphones and we sing and play and roast meat and have fun. (ID 70)*<br>*We'd do things like play a Korean traditional board game with the family during the holidays. (ID 79)* | ☐ Content: Planning and describing activities with family (language, executive function) |

## 4. DISCUSSION

This study examined the range of basic competencies required for delivering a cognitive enhancement program to older adults with MCI in urban and rural communities, identified their needs and memorable experiences, and evaluated the most appropriate content, delivery methods, and key intervention strategies. Based on these findings, we discuss important considerations for developing and implementing community-based programs.

### Basic competencies of older adults with MCI and strategies for program development

Previous research indicates that education affects cognitive abilities in older adults (Evans et al., 2019) and formal schooling contributes to cognitive reserve (Zahodne et al., 2015). However, this study involving older adults with MCI from urban and rural areas revealed no regional differences in cognitive function or health literacy. This finding suggests that, while educational disparities exist, they may not be the primary factor influencing cognitive performance. Instead, health literacy should also be taken into consideration. Therefore, cognitive enhancement program content for older adults with MCI should be simple, clear, and easily understandable to effectively improve health literacy.

There is a clear need for both group-based and individual home-based cognitive enhancement programs. Participants should be allowed to work at their own pace without feeling rushed. Approximately one-third of the older adults with MCI in this study lived alone; these individuals may face unique challenges, including heightened physical and mental vulnerability, limited nearby assistance, and sensory system deterioration (Hossain et al., 2022). These issues can result in poor adherence to cognitive enhancement programs, subsequently reducing self-confidence. To address this, strategies such as workbooks featuring repetition of content, large print, contrasting colors, and the use of microphones to amplify instruction can enhance participants’ self-confidence (Schmitter-Edgecombe et al., 2022).

Exercise improves cognitive function and helps prevent dementia in older adults (Xu et al., 2023). Participants in this study demonstrated good basic physical capabilities. However, over 90% had at least one medical condition, a quarter required a cane, and nearly half suffered from arthritis. Thus, exercise interventions must consider physical capacity limitations. Although the beneficial role of physical activity programs for older adults with MCI is widely recognized, fewer than half of the participants regularly engaged in exercise, with even fewer undertaking muscle-strengthening activities. A lack of facilities, transport availability, relevant services, and social engagement opportunities make engaging in physical activity particularly challenging in rural areas (Hancock et al., 2019; Pelletier et al., 2022). Digital and remote exercise programs could potentially increase physical activity among older adults with MCI in both urban and rural areas, despite these barriers (Prince et al., 2021).

Older adults own many mobile devices but often demonstrate low digital proficiency (Jun, 2021). The high smartphone ownership rate identified among older adults with MCI in this study aligns with similar findings from Korea (Korea Communications Commission, 2023), the United States (Pew Research Center, 2022), and the EU (Eurostat, 2022). However, there remains a gap between daily device access and effective usage (Jakobsson et al., 2019), with significantly lower proficiency among rural older adults compared to the general older populations in the US (Roque & Boot, 2018) and Japan (Shimokihara et al., 2024). Furthermore, cognitive limitations can negatively impact the ability to recall the procedural steps necessary for effective device use (Jakobsson et al., 2019). Physical impairments can also decrease precision and timing in touch interactions, while cognitive deficits may further hinder memory and device interaction. Digital interventions for older adults with MCI should feature understandable icons, voice commands, and simplified interfaces to minimize complex operations (Stara et al., 2023; De Santis et al., 2023).

### Needs and experiences of older adults with MCI and strategies for cognitive enhancement program development

To facilitate consistent participation of older adults with MCI in cognitive enhancement programs, it is crucial to first identify their particular needs for engaging with such programs. Our study has elucidated the underlying reasons for their reluctance, which include physical and emotional challenges. These findings are consistent with previous studies, which identified self-efficacy as a predictor of social participation (Hosseingholizadeh et al., 2019). Digital devices enable self-directed activities within the home environment, thereby facilitating participation by older adults with MCI who are reluctant to engage in external programs. Furthermore, when delivering programs, addressing participants by their preferred nicknames rather than by their actual names has been shown to help preserve dignity (Kim et al., 2023; Zhang et al., 2024). This approach not only safeguards participants’ personal space but also reduces the risk of stigma.

The objective of this study was to structure content around past experiences that individuals with mild cognitive impairment (MCI) would like to share, as past events represent familiar topics, and participants typically feel comfortable discussing memories from earlier life stages (Nieto-Vieites et al., 2024). In this study, participants were asked to identify significant memories they wished to recall. It was evident that memories considered most significant related to family, recreational activities, career, academics, and traditional Korean holidays.

This finding aligns with experiences reported by older adults in Europe, who identified family, leisure time, and experiences in nature as the most meaningful areas of their lives (Man-Ging et al., 2019). This observation can be interpreted as reflecting the integration of personal life within the broader life cycle, suggesting the importance of finding meaning through connections with oneself and others (Hupkens et al., 2018). However, cultural differences influence the key factors valued by older adults with MCI in various countries (Zhai et al., 2022). Consequently, when developing cognitive enhancement programs for older adults with MCI, it is advisable to include culturally sensitive content along with other key components, such as family and leisure activities.

Based upon the categories and sub-categories that emerged from interviews conducted in this study, the program’s content and delivery methods might be as follows: for individuals diagnosed with MCI, family is expressed not only through happy memories from the past but also in pride about having helped family members overcome adversity. The program content could incorporate reflective life story activities, expressed through writing or pictures, which potentially strengthen executive function (Moro et al., 2015). With regard to activities and play, the program could present games from a nostalgic perspective. This approach would evoke fond childhood memories, potentially fostering engagement in a variety of supplementary activities—such as crafts, physical exercise, and gardening—in addition to the main program.

In the section concerning career and academics, the subjects conveyed a blend of accomplishments in their professional pursuits and academic endeavors, alongside a sense of disappointment regarding their forced cessation of studies due to financial constraints during their formative years. Their aspirations for future learning and educational attainment were also highlighted. The provision of tangible acknowledgment, such as certificates of completion for program participation, name badges to facilitate learning experiences, and workbooks, as well as non-traditional reinforcement mechanisms such as praise stickers, is proposed to address the expressed needs and aspirations (Kim et al., 2023). Additionally, there is a direct demand for learning via digital devices, and therefore we recommend interventions that actively utilize these devices. Activities that are commonly enjoyed by older adults, such as food, clothing (hanbok), and games during traditional Korean holidays, can be connected to educational content incorporating various cognitive elements, including language, association, and calculation.

This study was conducted within a single urban and rural area of one country, which may pose limitations in generalizing the findings. Future studies should include a more diverse sample, encompassing different geographic regions and cultural contexts, to enhance the external validity and generalizability of the results.

## 5. CONCLUSION

This study investigated the basic competencies required to deliver a cognitive enhancement program for older adults with MCI in urban and rural communities. It identified the most appropriate content, delivery methods, and intervention strategies by assessing participants’ needs and memorable experiences. The cognitive enhancement program content should be specifically designed for older adults with MCI who have limited health literacy, and should incorporate exercise components. Program development is imperative and should include both group and individual sessions, conducted in the comfort and privacy of older adults’ residences. Programs should integrate life-cycle elements such as family, recreational activities, career, and academics, reflecting meaningful life contexts and cultural characteristics. These findings should be implemented to guide the creation of cognitive enhancement programs.

## Data Availability Statement

The data that support the findings of this study are available from the corresponding author upon reasonable request.

## Author contributions

Younkyoung Kim made substantial contributions to the study conception and design of this article, and acquisition, analysis, and interpretation of data. She drafted the article and revised it critically. Saeryun Kim contributed to the acquisition, analysis, and interpretation of data, as well as the drafting of the article. Asami Keiko contributed to the acquisition, analysis, and interpretation of data, as well as the critical revision of the article.

## Ethics approval statement

This study was approved by the Institutional Review Board of C University (South Korea) (IRB No. 1040198-220819-HR-104-02)

